# Paroxysmal Slow Waves Mark Ictal Networks

**DOI:** 10.64898/2025.12.22.25342212

**Authors:** Florent J. M. Boyer-Aymé, Hamza Imtiaz, Ofer Prager, Evyatar Swissa, Alaa Abu Ahmad, Yonatan Serlin, Refat Aboghazleh, Ilan Goldberg, Maayan Ben nun Caller, Idit Tamir, Oded Shor, Ben Whatley, Felix Benninger, Alon Friedman

## Abstract

Epilepsy diagnosis and treatment monitoring are hindered by the episodic, heterogeneous expression of seizures and by normal-appearing scalp EEG in many patients. We previously described paroxysmal slow-wave events (PSWEs)—brief epochs of broadband slowing detectable on EEG. Here, using intracerebral and epidural recordings in a paraoxon rat model of temporal lobe epilepsy, we show that PSWEs arise preferentially in temporo–frontal networks, co-occur with global slowing, and increase during both spontaneous and pharmacologically induced seizures.

Epidurally recorded PSWEs were temporally coupled to deep temporal discharges and were bidirectionally modulated by GABAergic agents (increased with pentylenetetrazol and decreased with pentobarbital).

In long-term video-EEG monitoring (LTM) patients with temporal lobe epilepsy, simultaneous stereo-EEG and scalp EEG showed that scalp PSWEs mirrored hippocampal spike-and-wave activity and were more often observed in the preictal and ictal periods than during interictal baseline.

These data indicate that surface PSWEs can index remote epileptiform activity and support their use as a quantitative, noninvasive biomarker for detecting EEG-silent deep foci and for pharmacodynamic.

## Introduction

Epilepsy stands out as one of the most prevalent chronic brain disorders, characterized by the susceptibility of cortical regions to generate seizures.^1^ The diagnosis of epilepsy remains challenging due to the transient nature of seizures, and the heterogeneity in their source and clinical presentation.^2–5^ In addition, the monitoring response to pharmacological, neuromodulatory or surgical therapies currently relies predominantly on patient reports that lack an easily accessible objective measure for their effect on brain epileptiform activity. Currently, electroencephalography (EEG) remains a gold standard for epilepsy diagnosis.^6^ However, assessment relies heavily on visual examination of EEGs to detect abnormal activity, such as seizures and inter-ictal epileptiform discharges (IEDs).^7^ This approach is limited, as EEGs may appear normal in up to 50% of epilepsy patients.^8^ IEDs can also be present in 10–12% of individuals without epilepsy,^8–11^ further complicating the accuracy of clinical diagnosis. Consequently, the development of diagnostic and pharmacodynamic biomarkers using quantitative EEG characteristics of the epileptic brain is essential. Enhanced power in slow frequency bands (delta and theta) coupled with reduced power in faster bands (alpha and beta) has been observed in various brain disorders,^12–17^ including epilepsy.^13^ We recently reported an algorithm for the detection of transient slowing of cortical activity recorded in inter-ictal EEGs from patients with epilepsy,^18,19^ which we termed “paroxysmal slow wave events” (PSWEs). PSWEs are characterized by a median power frequency (MPF) of 1–6 Hz for ≥5 s (1–5 Hz for ≥10 s in rodents). PSWEs were shown to be a robust discriminator between patients (and rodents) with epilepsy and healthy individuals, with AUCs >0.8 in a receiver operating characteristic (ROC) analysis.^18^ We also showed that PSWEs often co-localized with BBB dysfunction and are more common in patients with drug-resistance epilepsy.^20^ Additionally, we identified high rates of PSWEs as a predictive marker for epilepsy in patients presenting with a first seizure.^21^ Higher percentage of time in PSWE and lower MPF during PSWE were also found as predictive biomarkers for posttraumatic epilepsy (PTE) in patients following severe traumatic brain injury (sTBI).^19^ EEG slowing in patients with epilepsy was reported as far back as 1946,^22^ and later described in additional studies,^23,24^ as well as in adjacent to the seizure onset zone.^25^ However, the relationship between polymorphic (in contrast to rhythmic) EEG slowing and epileptic activity remains poorly understood. Moreover, while slow waves have been postulated to represent ictal events,^24^ others correlated slow activity with a reduction in cerebral blood flow (CBF) or speculated that these events represent post-ictal surrounding inhibition^26,27^ involving the recruitment of large populations of inhibitory neurons.^28–30^ In the current study, using epidural and intracerebral recordings in a rat model of temporal lobe epilepsy (TLE), we aimed to explore the spatial and temporal characteristics of paroxysmal slowing and its association with epileptiform activity and seizures. To translate these preclinical findings, we analyzed a publicly available EEG database of temporal lobe epilepsy (the European Epilepsy Database)^31^ and evaluated simultaneous scalp and intracerebral EEG recordings from a patient with TLE.

## Materials and methods

### Experimental design

To examine the effect of epileptogenesis on the occurrence of PSWEs, we first recorded from rats both prior to and eight weeks after induction of status epilepticus (SE), as well as from sham-treated control animals (n = 4 per group; timeline shown in Supplementary Fig. S1A). Recordings began one week before SE induction and continued for two months (Fig. 1E; Supplementary Figs. S4). In a separate cohort of animals (n = 10; Fig. 1F–G, Fig. 2–5), electrodes were implanted 4–8 weeks after paraoxon exposure, at which time animals exhibited spontaneous seizures (see timeline in Supplementary Fig. S1B and ^32,33^). To minimize detection of sleep-related slowing, PSWE analysis was restricted to data acquired 3–8 hours after the onset of the dark cycle.

**Figure 1.**
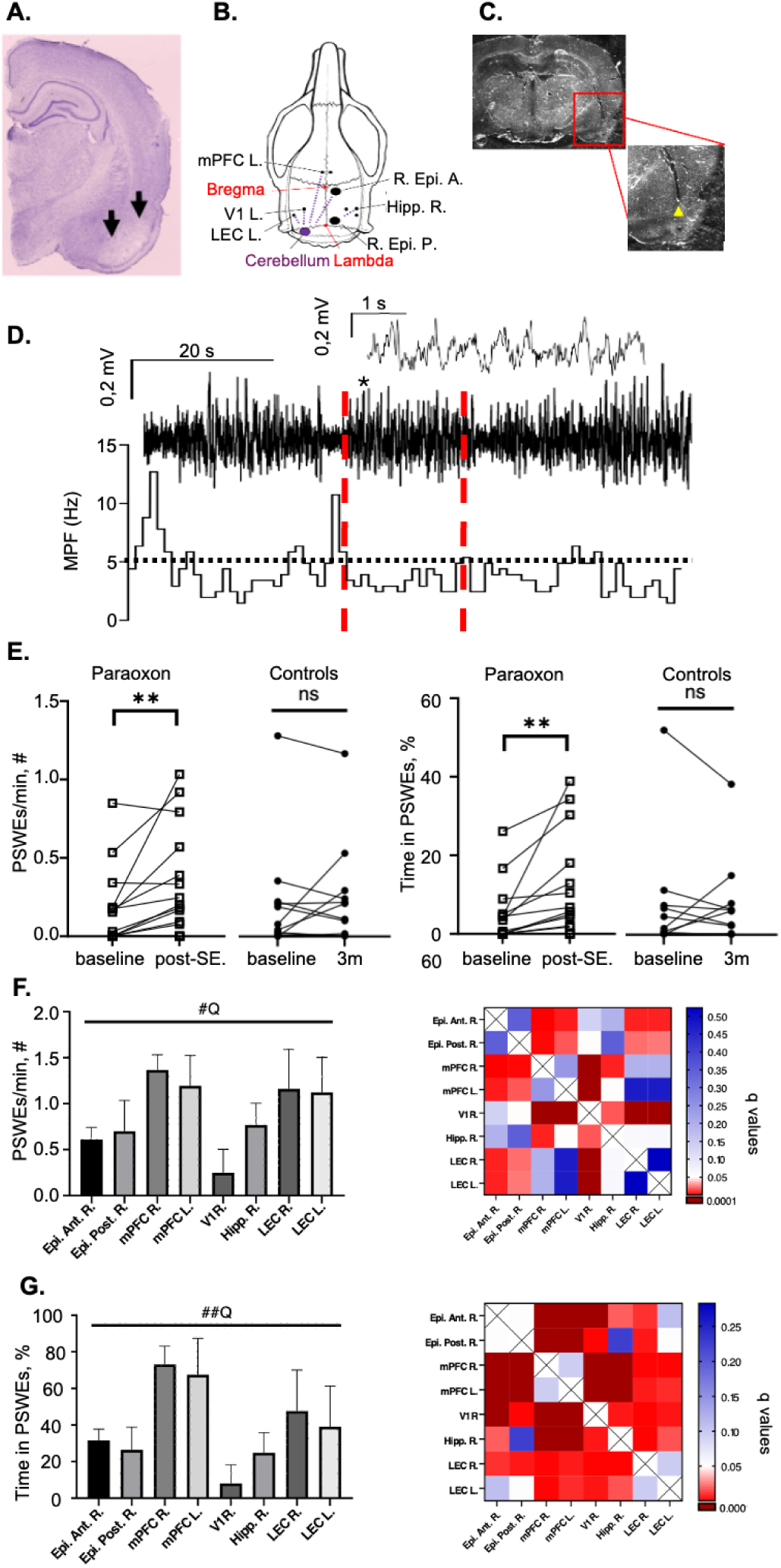
PSWEs in a rat model of temporal lobe epilepsy (TLE). **(A)** Cresyl-violet–stained coronal section 4 weeks after paraoxon-induced status epilepticus (SE) showing neuronal loss in the entorhinal cortex (arrows). **(B)** Electrode locations. **(C)** Coronal section from showing electrode tract in right lateral entorhinal cortex (LEC R.; yellow arrow). **(D)** Epidural recording: voltage (upper/middle trace) and median power frequency (MPF, lower trace). A PSWE is marked between red dashed lines; inset (upper trace) shows 5-s segment during a PSWE (asterisk). **(E)** PSWE features (pre-SE vs 3 months post-SE; epileptic rats, n = 16) vs sham controls (n = 15). **(F)** Left: PSWE occurrence (min⁻¹) in epileptic animals; Right: significance heatmap shows FDR-adjusted q-values (Benjamini–Hochberg correction of Kruskal–Wallis p-values for each brain region). **(G)** Left: percent time in PSWE in epileptic animals; Right: q-value heatmap (same correction family as in **F**). Red = q < 0.05 (significant); Blue = q ≥ 0.05 (not significant). Abbreviations: PSWE, paroxysmal slow-wave event; MPF, median-power frequency; LEC, lateral entorhinal cortex; SE, status epilepticus; R, right.

**Figure 2.**
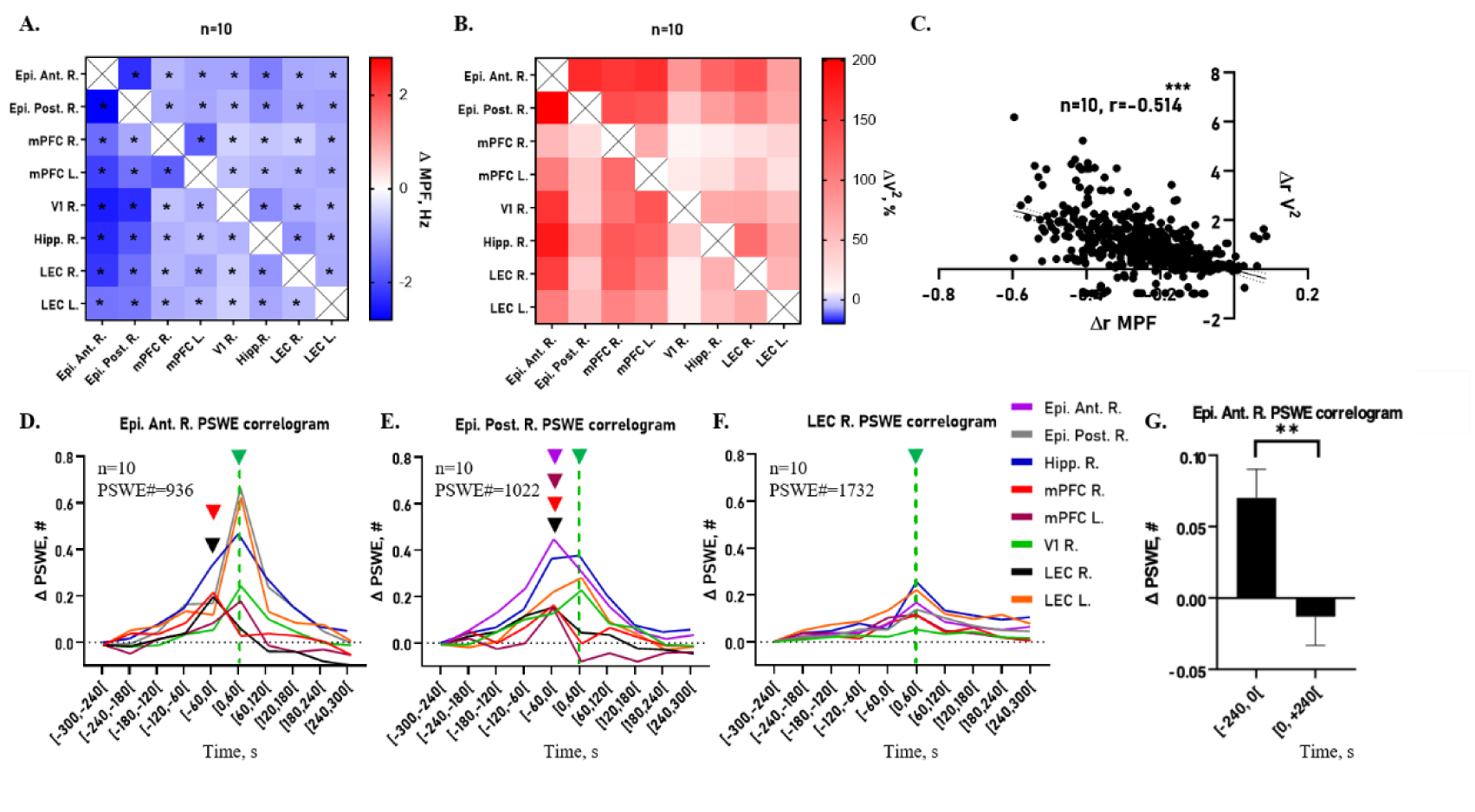
Inter-regional characteristics of PSWE occurrence in epileptic rats. **(A–B)** Changes in MPF (**A**) and power (V^2^; **B**) during PSWEs across recording sites. Rows indicate the reference channel with PSWE detection; columns show simultaneous values in neighboring channels. **(C)** Correlation between MPF and power changes across sites (Pearson, P < 0.0001), both expressed as ratios to baseline. **(D–F)** Correlograms aligned to PSWE onset in anterior epidural right (Epi. Ant. R.; **D**), posterior epidural right (Epi. Post. R.; **E**), and lateral entorhinal cortex right (LEC R.; **F**). PSWEs were counted in 60-s bins from −300 to +300 s, baseline-corrected to the [−300, −240[ s bin. The [0, 60[ s bin marks PSWE onset in the reference channel. Colored triangles (non-green) = peak PSWE rate peak PSWE occurence during or after the reference onset (see also Supplementary Fig. S3A–C). **(G)** Mean PSWE counts in LEC R. before [−240, 0[ s and after [0, 240[ s relative to baseline [−300, −240[ s during PSWEs in Epi. Ant. R. **Abbreviations**: PSWE, paroxysmal slow-wave event; MPF, median power frequency; LEC, lateral entorhinal cortex; Epi., epidural; Ant., anterior; Post., posterior; R., right.

**Figure 3:**
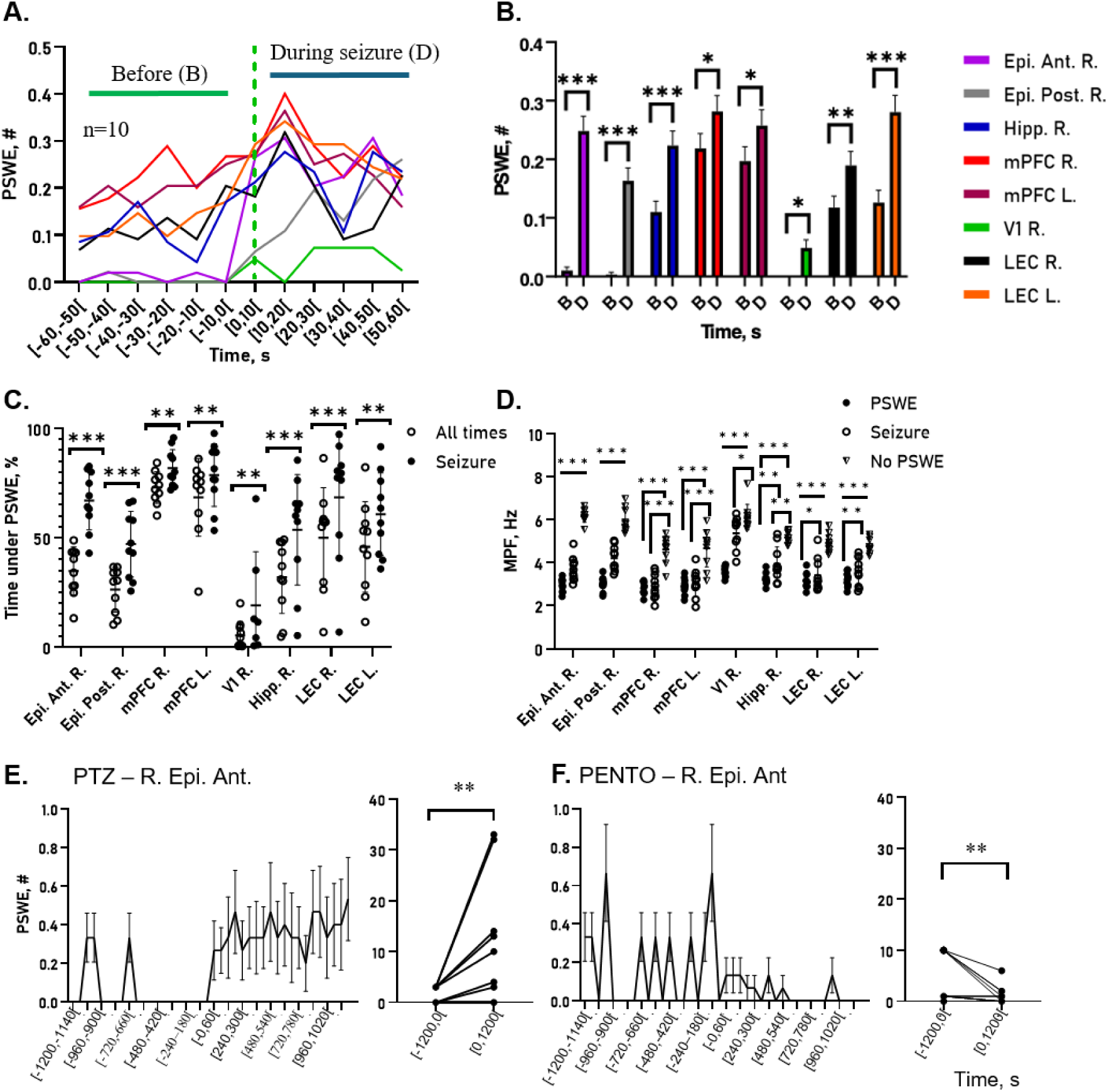
PSWE occurrence increases during seizures. **(A)** Correlogram of PSWEs and seizures within the same channel. PSWEs were counted in 10-s bins from -60 s to +60 s; t=0 marks seizure onset (green line). (**B)** Mean PSWE counts before [-60s, 0s[ and after [0s, +60s[ seizure onset across channels. **(C)** Percent time in PSWE during entire recording versus during seizures. **(D)** MPF during PSWEs, seizures, and seizures/ PSWE-free segments. **(E–F)** Effect of pentylenetetrazol (E) and pentobarbital (F) on PSWE occurrence.

**Figure 4.**
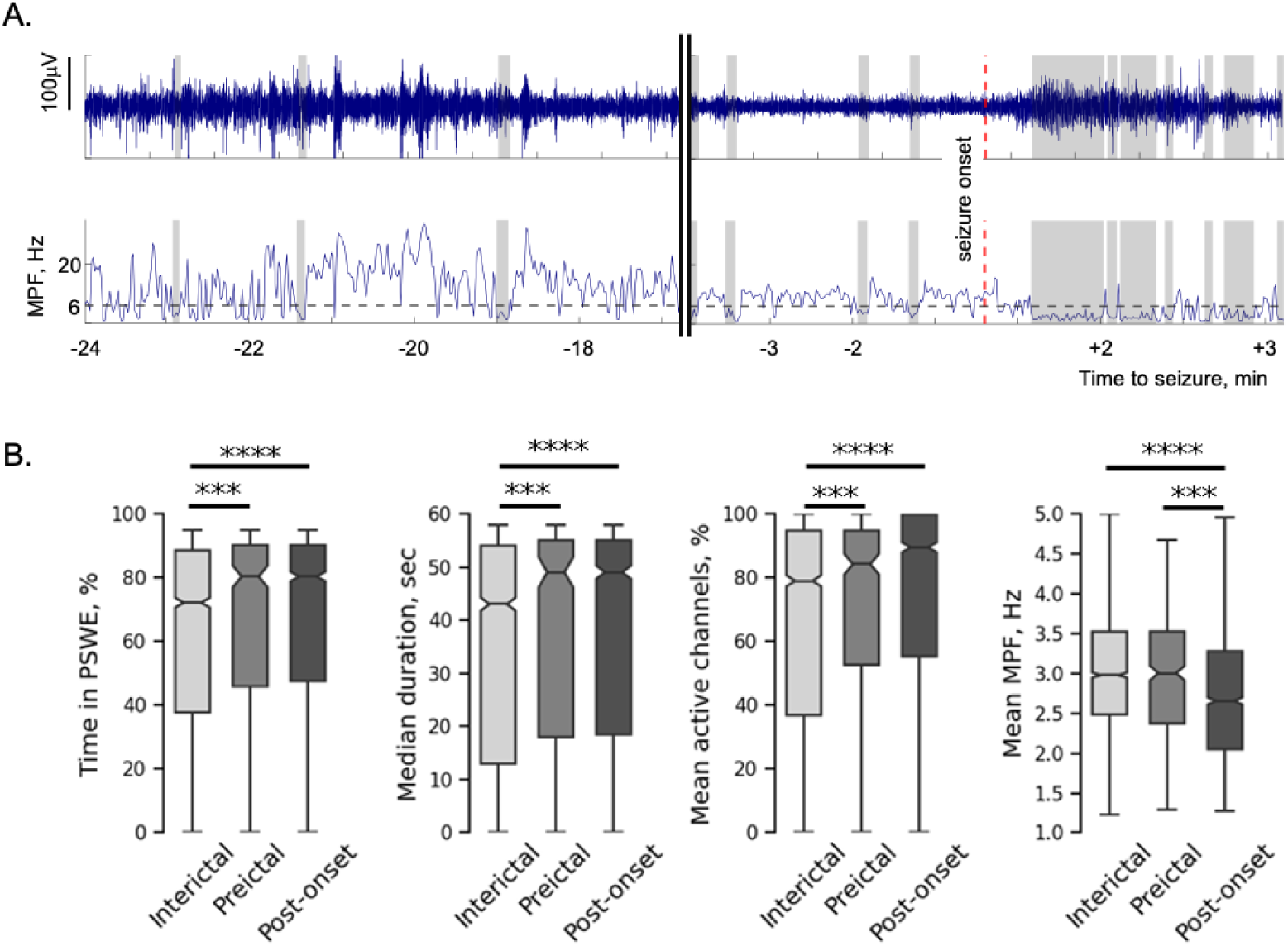
PSWEs during long-term scalp EEG in temporal-lobe epilepsy. **(A)** Example from T5 (left posterior temporal) electrode. Upper: voltage; lower: MPF. PSWEs were automatically detected when MPF was 1–6 Hz for ≥5 s (gray dashed line = 6 Hz threshold; shaded = PSWEs). Left: 7-min segment beginning 24 min before seizure onset. Right: 6-min segment spanning ∼3 min before and after seizure onset. **(B)** Quantification across seizures. Time in PSWE and PSWE duration were greater pre- and post-onset vs interictal. The number of simultaneously active channels was highest post-onset. MPF within PSWEs was lower post-onset than inter- and pre-onset. *** P < 0.001; **** P < 0.0001.

**Figure 5.**
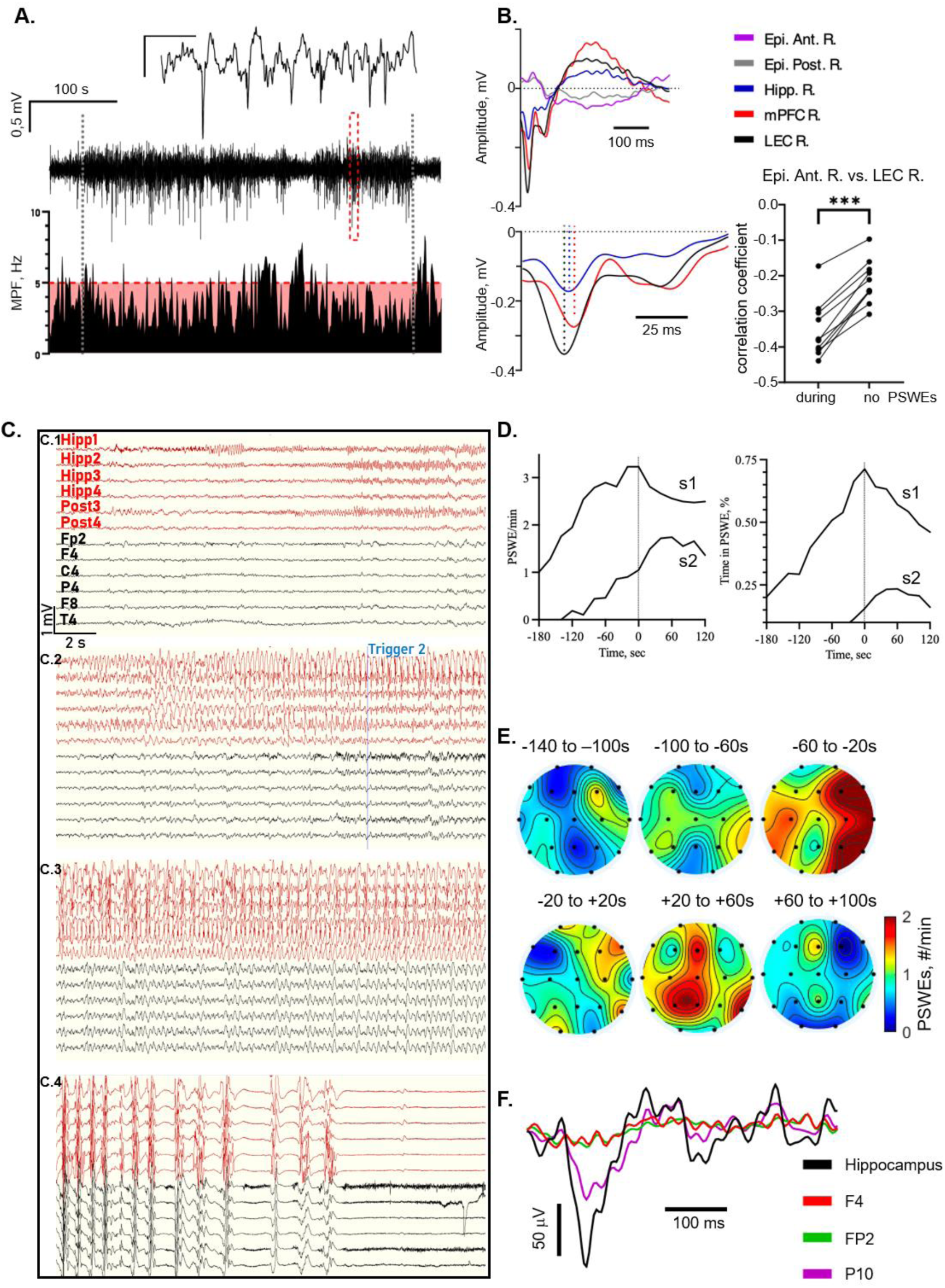
Epidural slowing accompanies remote deep epileptiform activity. **(A)** Rat example: seizure in entorhinal cortex with corresponding epidural MPF; red band marks PSWE range (1**–**5 Hz). **(B)** Average of 22 slow-wave patterns: spike peak appears in LEC R., then hippocampus (Hipp. R.), then mPFC R. Right: correlation between Epi. Ant. R. and LEC R. shows stronger negative coupling during PSWEs vs non-PSWE periods. **(C)** Patient with right TLE (long-term monitoring): hippocampal low-voltage fast activity preceeds clinical onset (“Trigger”, blue); surface EEG shows simultaneous slowing. Seizure evolved to bilateral tonic–clonic with post-ictal suppression. **(D)** PSWE occurrence (min⁻¹) and percent time binned around seizure onset for two seizures (s1, s2) show pre-ictal increases. **(E)** Scalp topography across four seizures shows higher PSWE occurrence in right temporal electrodes −60 to −20 s before onset. **(F)** Average of five hippocampal slow-wave events (Hipp1) aligned with slowing in scalp electrodes (F4, Fp2, P10, P4). ***Abbreviations***: PSWE, paroxysmal slow-wave event; MPF, median-power frequency; LEC, lateral entorhinal cortex; Hipp, hippocampus; mPFC, medial prefrontal cortex; Epi. Ant. R., right anterior epidural; EEG, electroencephalography; LTM, long-term monitoring; TLE, temporal lobe epilepsy.

### Ethics and animal care

All experimental procedures in animals were approved by the Animal Care and Use Ethical Committees at the Ben-Gurion University of the Negev, and were conducted in adherence to the NIH Guide for the Care Use of Laboratory Animals in Research.

### TLE model

Paraoxon-induced status epilepticus (SE) was used to trigger TLE, as reported.^32,33^ In short, Sprague Dawley rats (9–10 weeks old, 300–350gr body weight, provided by Harlan in Israel) were treated with paraoxon (Merck, USA) [intramuscular injections (IMs) of 0.45 mg/kg, equivalent to a 1.4 median lethal dose LD50 (lethal dose, 50% mortality when no further treatment is provided)] dissolved in propylene glycol: saline at a ratio of 1:27.25 (v/v). One min after paraoxon injection, animals were injected with atropine (Fluka, USA) (IM, 3 mg/kg, in saline) and toxogonin (Merck Serono, Israel) (IM, 20 mg/kg, in saline) to reduce the peripheral effects of the paraoxon and to prevent mortality. Midazolam (Rafa Laboratories, Israel) was administered (IM, 1mg/kg) 30 min following paraoxon exposure.

### Electrode implantation in animals

Rats were deeply anesthetized using a constant flow of isoflurane (2%) and oxygen; Body temperature was maintained throughout the procedure at 37°±0.2°C using a heating pad (Mirom Medical & Research Equipment LTD, Israel). Thin epidural (screw diameter, 1000 μm, Palboreg, Israel) and intracerebral silver (coated diameter, 203.2 μm, AM-system, USA)/tungsten (coated diameter, 50.8 μm, WireTronic Inc., USA) wire electrodes were implanted using a digital stereotaxic instrument (RWD LifeScience). In a subgroup of animals, tungsten electrodes were used. Intracerebral electrodes included the dorsal peduncular (DP) cortex, the most ventral part of the medial prefrontal cortex (mPFC) bilaterally, CA1 regions of the right hippocampus (Hipp), right basolateral amygdala (BLA), bilateral lateral entorhinal cortex (LEC) and visual cortex (V1). One anterior and one posterior epidural electrode were also implanted on the right side of the skull. The anterior electrode (Epi. Ant. R.) was inserted above the primary (M1) and secondary (M2) motor areas, while the posterior electrode (Epi. Post. R.) recorded signals in the retrosplenial cortex (RSA), as well as in the posteriomedial (PM) and primary (V1) visual areas. A reference was placed over the cerebellum and a ground electrode was inserted in the neck (for the coordinates used, see Supplementary Table 1). Following implantation, animals were returned to their cage, and allowed to recover for 7–14 days before recordings started. Emphasis was given to temporal lobe regions (Hipp, BLA, LEC), considering their association with the ictal onset zone and the described histopathology.^34^

### Histology

At the end of the recording period, rats were anesthetized with ketamine/xylazine (i.p. 79.2 and 59.4 mg/kg), followed by intracardiac perfusion with phosphate-buffered saline (PBS) and 4% paraformaldehyde (PFA). Brains were removed, post-fixed, and transferred sequentially into 10%, 20% and 30% sucrose-PBS (24-72 h each) before freezing and sectioning (20–40 μm using a cryostat). Cresyl-violet staining^35^ was used to assess damage and verify electrode placement under a light microscope (Axioskope 2plus, Zeiss, Germany).

### Signal recording and analysis in animals

Data were acquired using rodentPACK (Emka Technologies, France) or PowerLab 8/35 (ADInstruments, New Zealand) at 1000 Hz and band-pass filtered (1–45 Hz). PSWEs were detected using custom MATLAB scripts when the median power frequency (MPF) was 1–5 Hz for ≥10 s, as described.^18,36^ Metrics included PSWE occurrence (min^-1^), time in PSWE (% of recorded time with PSWEs), mean duration, and MPF. Seizures and interictal activity were identified by visual inspection (LabChart 8, ADInstruments).

### MPF and power variation

To test if PSWEs in one channel were associated with remote activity changes, MPF was calculated in all channels during PSWE and non-PSWE periods. MPF variation was defined as:

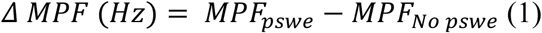

Where MPF*_pswe_* and MPF_No *pswe*_ are the mean MPFs in the “adjacent” channel during PSWE and non-PSWE activity in the “reference” channel, respectively. Power was estimated as mean squared amplitude [*V*^2^], within each segment and percentage change was calculated as:

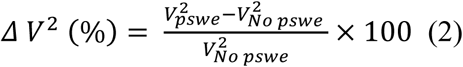

For correlation analysis MPF and power variation were calculated as ratios:

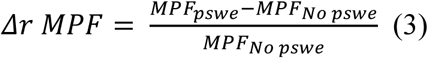

for the MPF and

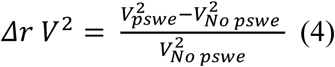

for the power.

### Correlogram analysis

Correlogramd were computed with custom MATLAB scripts to examine temporal relationships between PSWEs in different regions, and between seizures and PSWEs. For each PSWE occurring in a reference channel, PSWEs in other channels were counted in 60-s bins, and rates were normalized to baseline (-300 s and -240 s). For PSWEs vs seizure correlograms, seizures with a clear onset (>60 s duration) were identified visually (LabChart), and PSWEs were counted within 10-s bins around onset.

### Low-frequency correlation analysis

To assess coupling between surface and deep electrodes during PSWEs, recordings from epidural (Epi. Ant. R.) and entorhinal cortex (LEC R.) electrodes were band-pass filtered (1–5 Hz). Correlations were computed (MATLAB corrcoef) during epidural PSWEs versus non-PSWEs period.

### Pharmacological experiments

These experiments were performed in paraoxon-treated rats with established epilepsy (8–12 weeks post-injection). Animals recieved i.p. injection of the pro-convulsant GABA_A_ receptor antagonist, pentylenetetrazol (10mg/kg) or the anti-convulsant barbiturate pentobarbital (4mg/kg), five times at 30 min interval. Intracerebral activity was recorded before and during treatment to assess effects of GABAergic modulation on PSWEs. Animals were killed 20 min after the last injection.

### Human recordings and analysis

The retrospective study of anonymized patient data was approved by the Rabin Medical Center IRB. Stereoencephalography (sEEG) with simultaneous scalp EEG was performed during routine pre-surgical evaluation. Five depth macroelectrodes (8–10 contacts, 5-mm spacing; Ad-Tech, USA) were stereotactically implanted (CRW frame, Integra, USA) targeting the amygdala, anterior and posterior hippocampus, posterior temporal, and occipitotemporal cortex. Planning was based on pre-operative 3T MRI (Philips, Netherlands) co-registered with intraoperative CT (O-arm, Medtronic, Ireland) using Stealth 8 navigation (Medtronic); post-operative CT confirmed electrode placement. Recordings began 24 h post-surgery, sampled at 2 kHz and downsampled to 250 Hz. Scalp EEG (10–20 system, 19 electrodes) was sampled at 250 Hz. Patients pressed a trigger button at seizure onset.

Data were preprocessed in EEGLAB^37^ as reported ^18,19,21^: band-pass filtered (1–45 Hz), decomposed with ICA, and artifact components (muscle, eye, heart) removed using ICLabel.^38^ Preprocessed EEG was segmented into 2-s windows (50% overlap) and MPF calculated. PSWEs were defined as MPF 1–6 Hz for ≥5 s in ≥2 electrodes.

For scalp EEG during long-term monitoring, data were stored in 1-h blocks (University of Freiburg SQL servers). The block containing a seizure and the preceding hour were retrieved, merged, and aligned to seizure onset, yielding 60 min preictal and 20 min postictal data. Recordings with ictal activity in the 1-hour period preceding the seizure onset or data gaps > 3 seconds were excluded.

### Statistical analysis

Analyses were performed in GraphPad Prism 9. When PSWE features (occurrence, duration, MPF) failed normality, data were analyzed with the Kruskal–Wallis test; normally distributed measures (% time in PSWE, MPF during PSWE) were tested with mixed-effects ANOVA (Geisser–Greenhouse correction). Correlogram data were analyzed with non-parametric tests. False discovery rate (FDR) correction (two-stage step-up, Benjamini, Krieger, and Yekutieli) was applied to ANOVA, Kruskal–Wallis, Friedman, and 2-way ANOVA. Q values (q) present FDR-adjusted significance with qmin/pmin = strongest significance and pmax/qmax = weakest.

## Results

### PSWE occurrence in epileptic animals differs across brain regions

Consistent with prior results,^18^ PSWE occurrence, duration, and time in PSWEs increased in epileptic rats versus pre-SE baseline (Fig. 1E; Supplementary Fig. S2A), with no change in controls. PSWEs were detected across all electrodes but occurred more frequently in the right medial prefrontal cortex (mPFC R.) and lateral entorhinal cortex (LEC R.) than in the visual cortex (V1 R.) (1.37±0.16, 1.16±0.43 and 0.25±0.25 PSWE/min, respectively, P < 0.0001, ANOVA; Fig. 1F). The percent time in PSWEs was also higher in the mPFC R. and LEC R. than V1 R. (73.14±10%, 47.6±22.5% and 8.01±10.17%, respectively, P < 0.0005 ANOVA; Fig. 1G). Similarly, PSWE duration was longer in mPFC R. (32.7±6.7s) and LEC R. (24.1±8.3s) than V1 R. (15.02±5.17s, P < 0.03; Supplementary Fig. S2C). The MPF was lower in mPFC R. ((2.85±0.3) but not LEC R. (3.18±0.31) compared to V1 R. (3.55±0.4; P = 0.005; Supplementary Fig. S2D). These findings indicare region-specific PSWE patterns. For ANOVA details, see matrices (#Q, ##Q; Fig. 1F, G).

### PSWEs are associated with global slowing

To test whether PSWE in one region was linked to remote changes, we measured MPF across channels during PSWEs recorded in any given channel. PSWEs in one channel were associated with a slowing (MPF reduction) in others (-1.22 ± 0.8 Hz; Fig. 2A; two-way ANOVA, Pmax = 0.0217, Pmin < 0.0001), and with increased power (94.32 ± 102.5%; Fig. 2B). MPF reduction correlated with power increase (r=-0.514, P < 0.0001, Pearson correlation; Fig. 2C), suggesting greater network synchronization during PSWEs. Correlogram analysis showed that PSWE rate rose in mPFC R. and LEC R. 60–1 s before detection in the anterior epidural electrode (Fig. 2D,G; qmax = 0.0040, qmin < 0.0001, and qmax = 0.063 and qmin < 0.0001, Friedman test for the mPFC R. and LEC R., respectively). PSWEs in the posterior epidural electrode appeared with a delay relative to the anterior epidural electrode (Fig. 2E), indicating propagation. Across channels, PSWE peaks coincided with or followed onset in LEC R. (Fig. 2F–G). These findings suggest that PSWEs preferentially involve ictogenic regions and induce slowing (see Discussion).

### Occurrence of PSWE increases during seizures

Spontaneous seizures were recorded in all paraoxon-treated rats. Video-EEG showed behavioral arrest with or without jerks or tonic seizures. Correlograms analysis revealed that PSWE occurrence was higher during seizures than in pre-ictal period (-60 to 0 s; Fig. 3A-B; two-way ANOVA, qmax = 0.0315, qmin < 0.0001). Across channels, time in PSWE was greater during seizures compared to overall baseline (Fig. 3C, qmax = 0.0067, qmin < 0.0001). MPF was the lowest during a PSWE, higher during seizures, and highest outside PSWEs (Fig. 3D).

To test whether PSWEs reflect hyperexcitable state or more inhibitory drive, epileptic rats recieved the pro-convulsant GABAergic antagonist pentylenetetrazol (PTZ) or positive GABA_A_ modulator pentobarbitol (PENTO). PTZ increased PSWE occurrence and triggered epileptiform activity and seizures (2/3 rats after one injection; one rat developed SE after the fifth; Fig. 3E). Pentobarbial reduced PSWE rates and spontaneous movement (Fig. 3F). These findings suggest that PSWEs reflect hyperexcitable network states suppressed by GABA_A_ transmission.

### PSWEs in human EEG recordings

To examine PSWEs in human patients, we analyzed retrospective EEG data from 18 individuals with temporal lobe epilepsy (137 seizures; European database, see Methods). PSWEs were detected during interictal, preictal (∼3 min) and post-onset periods (Fig. 4A). PSWE occurrence, duration, and multi-channel involvement were greater in the preictal and post-onset periods compared with interictal periods (-60 minutes to -3 min; Fig. 4B). PSWE median frequency was lower post-onset than during interictal and preictal epochs (Fig. 4B).

### PSWEs reflect deep epileptiform activity

Rodent and human data indicated that PSWEs are linked to network hyperexcitability and seizures. To test whether surface PSWEs reflect remote epileptiform activity, we averaged spike-wave (SW) discharges during seizures in rats (Fig. 5A). SWs consistently originated in the LEC followed by hippocampus and mPFC R. (Fig. 5B). Wave components were simultaneously observed in epidural electrodes with inverted polarity, but absent in V1 R. electrode (Supplementary Fig. S3D). Correlation between LEC R. and ipsilateral epidural signals was stronger and negative during PSWEs vs background (no PSWEs) (r=-0.35 vs. r=-0.21, n=10, P < 0.0001, paired t-test; Fig. 5B). These findings suggest that PSWEs reflect cortical slowing driven by deep epileptiform discharges.

We next analyzed simultaneous surface and sEEG recordings from a right-handed man in his 30’s with drug-resistant temporal lobe epilepsy (tumor in the right temporal lobe, extending to the temporal horn of the right lateral ventricle, involving the head and body of the hippocampus; Supplementary Fig. S5). Since seizure onset could not be localized using video EEG long-term monitoring, five sEEG electrodes were implanted in the right temporal lobe, specifically in the amygdala, anterior and posterior hippocampus, and posterior temporal and temporo-occipital junction. sEEG showed seizure onset in the posterior hippocampal electrode (contacts 1 and 2) with spread to the posterior temporal cortex (contact 1, Fig. 5C). Clinical onset (color distortion, staring, oral automatism) occurred 9 s later (indicated by “Trigger 2” in Fig. 5C.2), while EEG remained silent until spread. Across four seizures, PSWEs occurrence, % time and duration rose in right temporal electrodes 20–60 s before clinical onset and the appearance of the electrographic seizure on the scalp EEG (Fig. 5D-E). As in rats, hippocampal spike-waves were associated with surface slowing (Fig. 5F).

## Discussion

Recent work suggests that paroxysmal slow-wave events (PSWEs) may mark epileptic brain states.^18,21,36^ We show that PSWEs preferentially arise in temporo–frontal networks, increase preri-ictally, correlate with deep temporal discharges, and are modulated by GABAergic drugs—supporting their role as a noninvasive index of ictogenic activity.

In rats, PSWEs were most prominent in entorhinal cortex and ventral mPFC—regions implicated in TLE. Occurrence in one site coincided with global slowing and power increases, consistent with network synchronization. Correlogram analyses suggested initiation in anterior temporal structures with subsequent neocortical spread (Fig. 2D–G, Supplementary Fig. S3A–C). PSWEs rose during seizures, consistent with a transition to a hyperexcitable state (Fig. 3, Supplementary Fig. S4).

Mechanistically, several findings link PSWEs to excitatory drive from deep foci: (i) PSWEs increased during seizures and SE; (ii) PTZ (GABA_A_ antagonist) increased PSWEs whereas pentobarbital (positive GABA_A_ modulator) decreased them; (iii) in rats, hippocampal/entorhinal/mPFC spike–wave activity co-occurred with epidural slowing and stronger negative correlations between deep and surface signals during PSWEs; and (iv) in the sEEG/scalp case, hippocampal fast activity preceded clinical onset while scalp EEG showed synchronized slowing, with PSWEs increasing minutes before scalp ictal patterns emerged.

An inhibitory contribution to PSWEs generation cannot be excluded.^39^ GABA_A_-mediated transmission can promote seizure-like events in the rat hippocampus via back-propagation from CA3/hilus to the dentate gyrus.^40^ Hypersynchronous GABAergic-mediated transmission was also described in the BBB breakdown model of epileptogenesis, in which low dose application of the GABAergic antagonist bicuculline led to short-term inhibition of both spontaneous and evoked epileptic paroxysmal activity.^41^ Surround inhibition via interneurons activation and GABA_B_-mediated K^+^ conductance^28–30,42,43^ may also generate slow waves of inhibition, as seen during SW discharges.^39^ As such, the remote opening of GABA_B_ channels might be synchronized with increased Ca^2+^-dependent K^+^ conductance within the epileptic foci.^39^ Such surround inhibition could explain the remote hypersynchronized slowing we observed.

Overall, PSWEs likely capture a mixture of polymorphic slowing that includes—but is not limited to intermittent rhythmic delta activity (IRDA - 2–4 Hz).^39–42^ By design, our detector targets prolonged (≥5 s in humans; ≥10 s in rodents) 1–6 Hz events with notable theta content, a profile that previously separated patients with epilepsy from controls (AUC > 0.8). Together with the observation that PSWEs localization may predict BBB dysfunction^18^ and focal lesion,^44^ our data suggest the potential of PSWEs as a biomarker for atypical epileptiform activity generated within deep brain structures, specifically within the ictogenic zone.

Compared with previor studies,^18,36^ control rats showed relatively high PSWE rates, possibly reflecting electrode-related injury and known spike–wave susceptibility in Sprague–Dawley rats.^45^ Still, PSWEs rose with epileptogenesis and were higher in epileptic animals than controls, supporting disease specificity.

Translation to humans mirrored the animal findings: in 18 TLE patients (137 seizures), Time in PSWE, duration, and spread were greater pre- and post-onset vs interictal, with MPF reduction post-onset. In the sEEG/scalp case, hippocampal spikes temporally aligned with scalp slowing, and right temporal scalp PSWEs increased 20–60 s before clinical onset—suggesting diagnostic utility when scalp EEG appears normal.

Limitations include single sEEG case, possible drug/state confounds, lack of coverage of extra-temporal brain regions that may be important for propagation of epileptiform activity (e.g. thalamus),^46,47^ and uncertain generalizability to extratemporal epilepsies. Prospective, multi-center studies with source-localized PSWEs, thalamic recordings, and standardized pharmacological challenges are warranted, as is testing whether PSWEs can guide imaging in remote areas (specifically in low- and middle-income countries),^44^ presurgical planning or quantify pharmacodynamic response. Because PSWEs are also elevated in sub-group of patients with Alzheimer’s disease,^18^ they may also serve as markers of subclinical seizures in neurodegeneration.^48^

In sum, PSWEs provide a scalable, quantitative, noninvasive marker of deep epileptiform activity and seizure propensity. Incorporating PSWE metrics alongside traditional IED detection could improve diagnosis, identify EEG-silent deep foci, and offer a practical pharmacodynamic readout in trials.

**Figure 6.**
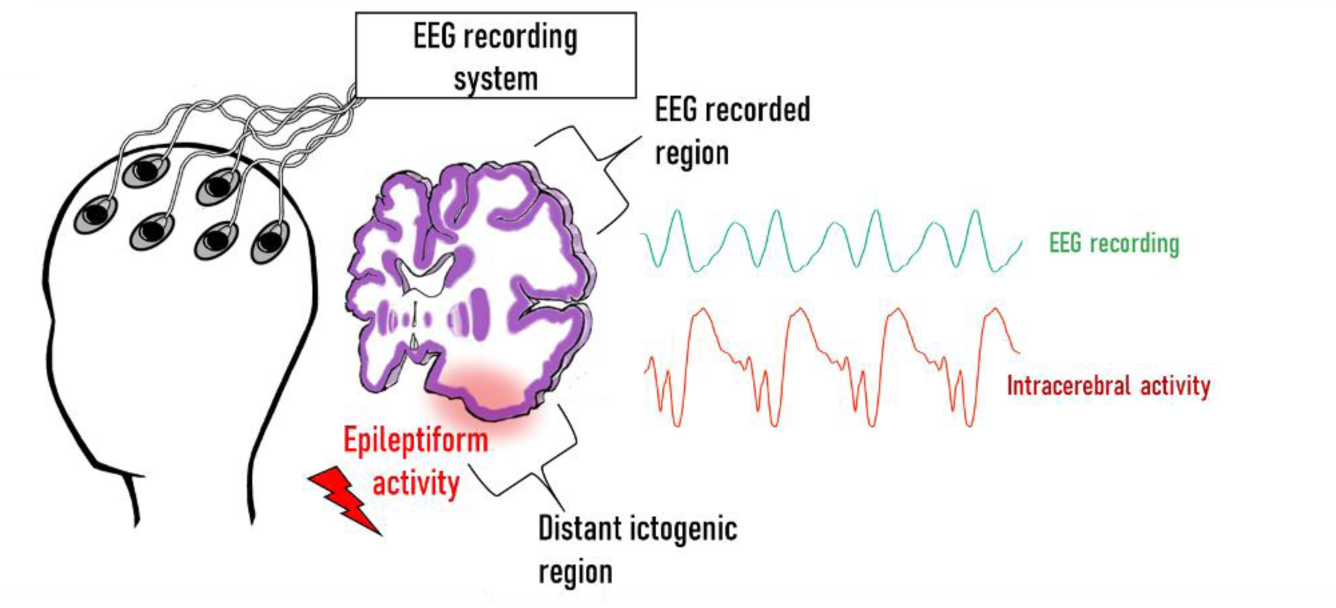
PSWEs index remote epileptiform activity (schematic). In temporal lobe epilepsy, scalp-recorded PSWEs reflect hypersynchronous discharges in deep/anterior temporal cortex, remote from recording electrodes. The PSWE algorithm captures epileptiform-related slowing, including OIRDA and TIRDA. **Abbreviations:** PSWE, paroxysmal slow-wave event; EEG, electroencephalography; OIRDA, occipital intermittent rhythmic delta activity; TIRDA, temporal intermittent rhythmic delta activity.

## Supporting information

Suplemental material - Florent Boyer Ayme 2026

## Acknowledgements

The authors would like to thank Dr. Tawfeeq Shekh-Ahmad for his advice on implant durability and Kay Murphy for her technical assistance.

## Data availability

All data needed to evaluate the conclusions in the paper are present in the paper and/or the Supplementary Materials. Scripts used in this manuscript will be made available on a suitable platform with a material transfer agreement.

## Author contributions

[AF] conceived the study and secured funding; [FBA] and [HI] developed the detection/analysis pipeline; [FBA], [RA], [OP], [ES]and [AAA] performed animal experiments; [AF], [FBA] and [HI] conducted formal analyses and created figures; [IG], [MBC], [IT], [OS] and [FB] coordinated clinical recruitment, and collected and analyzed EEG/sEEG data; [AF] provided supervision. [FBA], [YS], [BW] and [AF] drafted the manuscript; all authors revised the manuscript critically for important intellectual content and approved the final version.

