## Supplementary material for "Paroxysmal Slow Waves Mark Ictal Networks": Suplemental material - Florent Boyer Ayme 2026

Florent J. M. Boyer-Aymé et al.

**This PDF file includes:**

Figs. S1 to S5

Tables T1

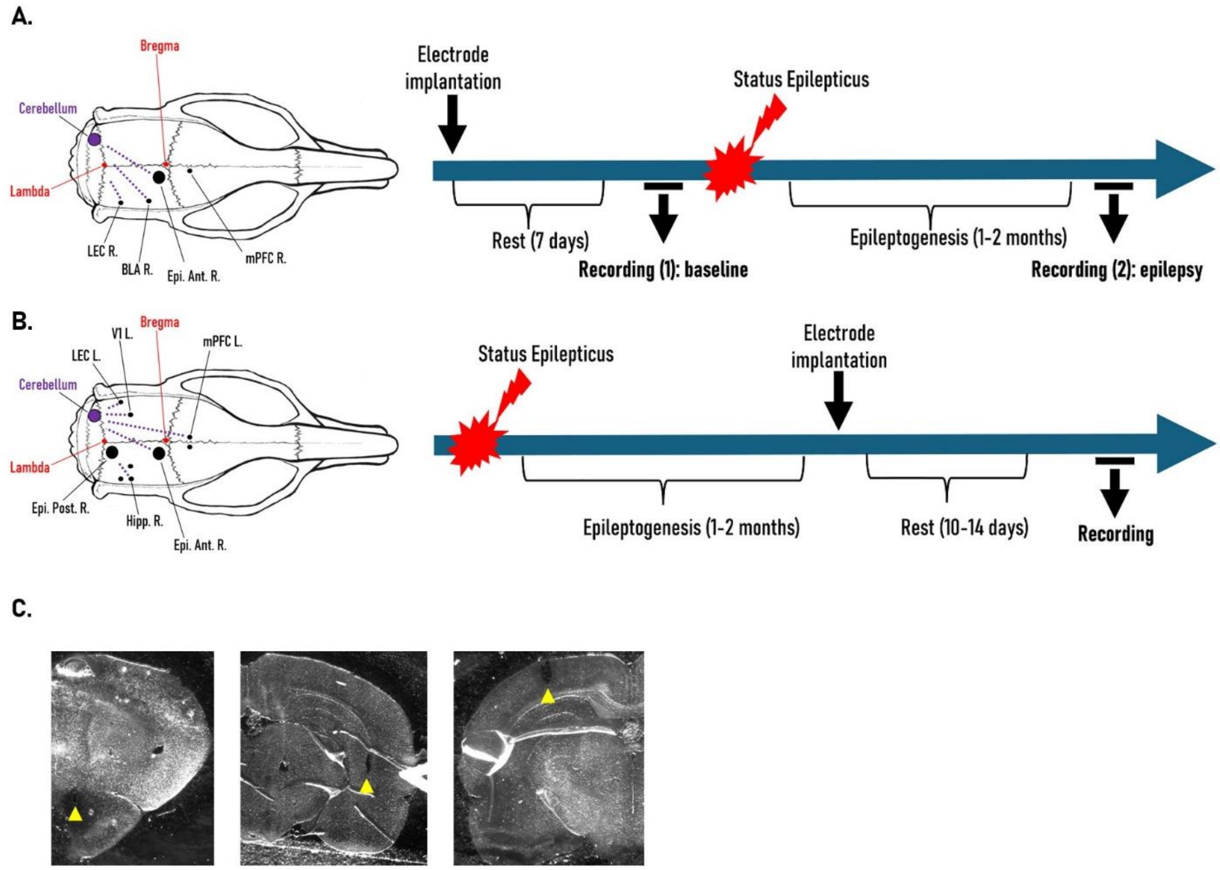


**Fig. S1. Experimental timelines, electrode montages, and histology.** (A) Montage and timeline for the epilepsy validation experiment (related to Fig. 1): right lateral entorhinal cortex (LEC R), right basolateral amygdala (BLA R), right medial prefrontal cortex (mPFC R), and right anterior epidural electrode (Epi. Ant. R). Status epilepticus (SE) was induced after electrode implantation to assess within-animal PSWE changes following epileptogenesis. (B) Montage and timeline used in most experiments (related to Fig. 1F–G, Fig. 2–5): bilateral LEC, visual cortex (V1), mPFC, right anterior and posterior epidural electrodes (Epi. Ant. R, Epi. Post. R), and right hippocampus (Hipp. R). (C) Coronal sections (bright-field) illustrating representative implant tracks in ventral mPFC, hippocampus, and V1; yellow arrow marks electrode tip.

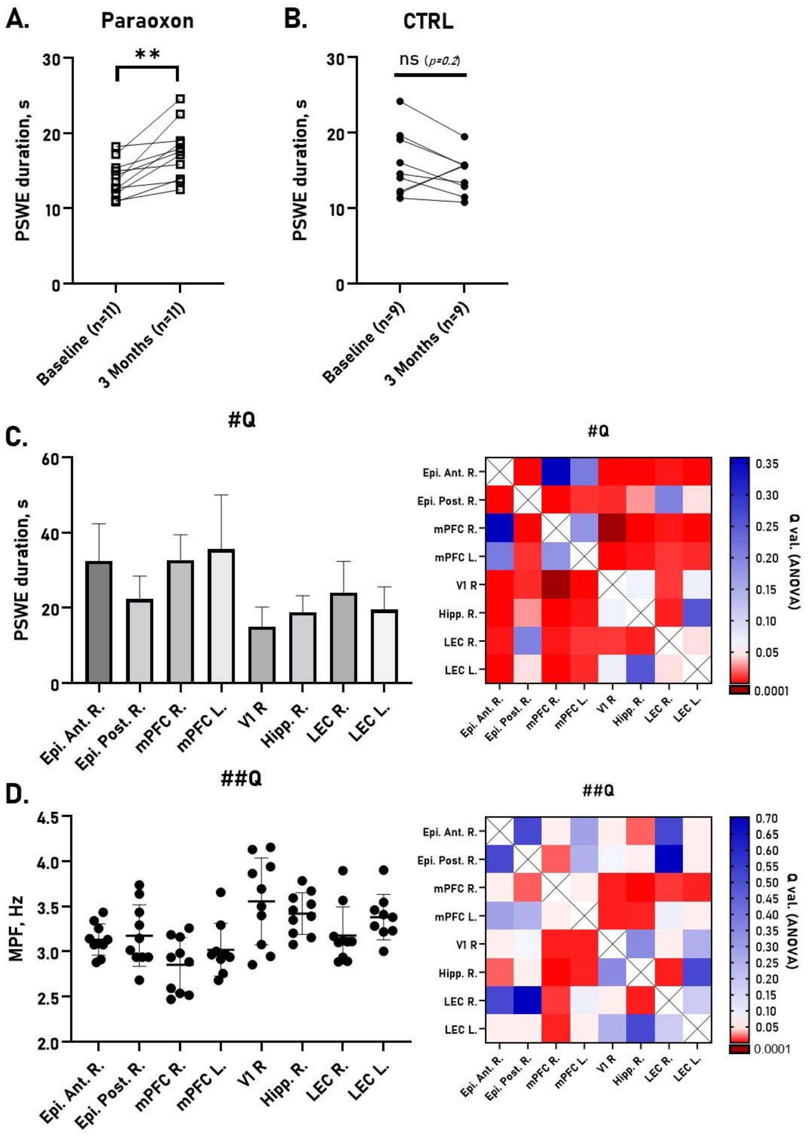


**Fig. S2. Brain regions differ in PSWE rate and spectral features.** (A,B) Validation experiment with channels pooled for statistics: PSWE duration increased only in paraoxon-poisoned animals (white squares) versus controls (black dots). (C,D) Unipolar recordings as in timeline Fig. S1B: heatmaps show significance (#Q, ##Q) from Kruskal–Wallis tests with Benjamini–Krieger–Yekutieli FDR correction; red, $q<0.05$; blue, $q>0.05$. (C) PSWE duration (s) varies across regions in poisoned epileptic rats. (D)

Median power frequency (MPF) during PSWEs varies across regions in the same animals.

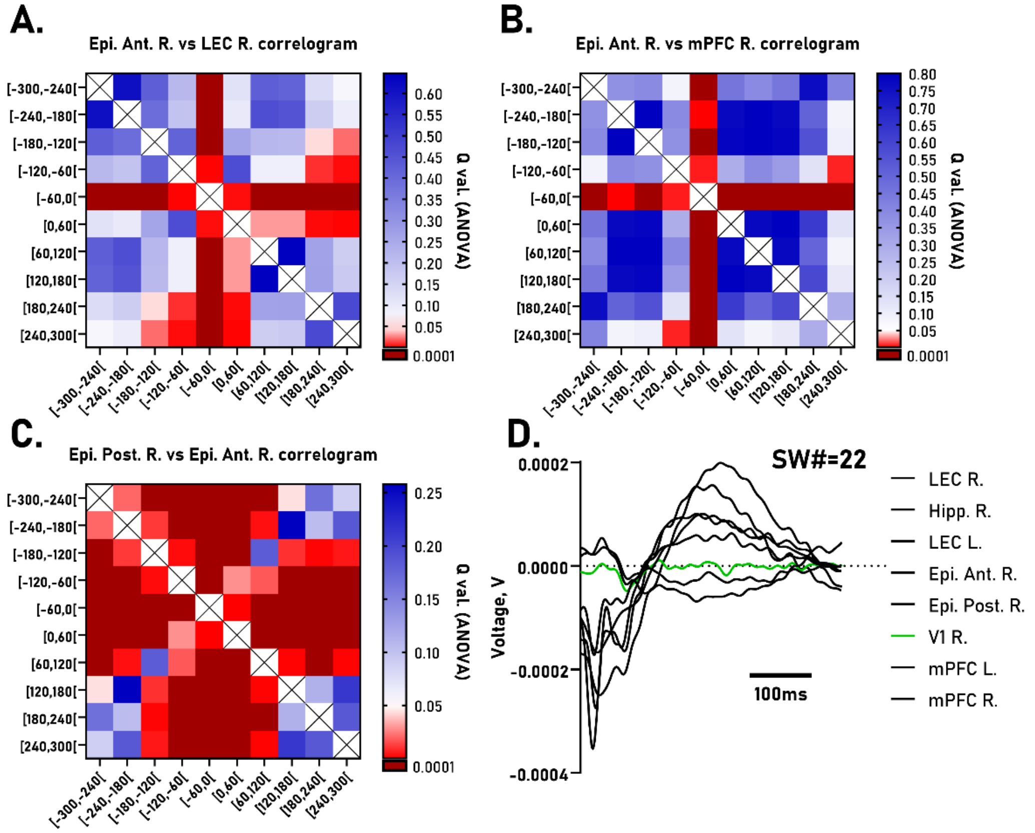


**Fig. S3. Statistics for PSWE–PSWE correlograms and lack of coupling with V1 activity.** (A) Epi. Ant. R (channel of interest) vs LEC R: the interval −60 to 0 s differs significantly from all other bins, indicating a surge of PSWEs in LEC R preceding Epi. Ant. R PSWEs ($q_{min}<0.0001$; $q_{max}=0.0063$). (B) Epi. Ant. R vs mPFC R shows a similar pre-event surge ($q_{min}<0.0001$; $q_{max}=0.015$). (C) Epi. Post. R vs Epi. Ant. R: a pre-event surge in Epi. Ant. R precedes Epi. Post. R PSWEs ($q_{min}<0.0001$; $q_{max}=0.0007$). Friedman test with Benjamini–Krieger–Yekutieli two-stage step-up FDR correction. (D) Full dataset for Fig. 5B including V1 shows that V1 activity does not correlate with slow-wave events in LEC R, Hipp. R, or mPFC R

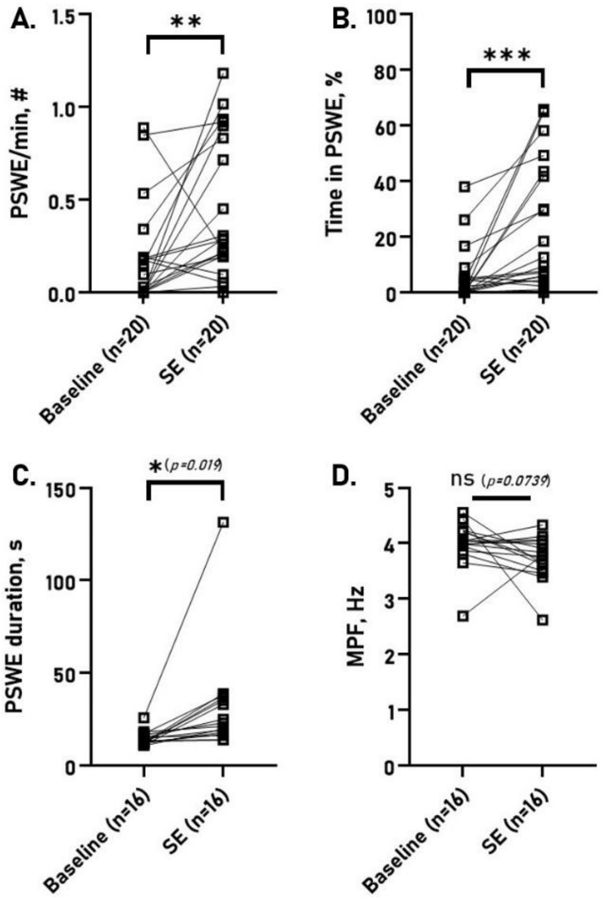


**Fig. S4. PSWE occurrence increases during status epilepticus.** Rats recorded during paraoxon-induced SE (n=5); channels pooled owing to sample size (as in Fig. 1E). Depending on normality, paired parametric or nonparametric tests were applied (see Methods). (A) PSWEs per minute increased during SE. (B) Percentage of time in PSWE increased during SE. (C) PSWE duration increased during SE. (D) No. significant change in MPF during SE.

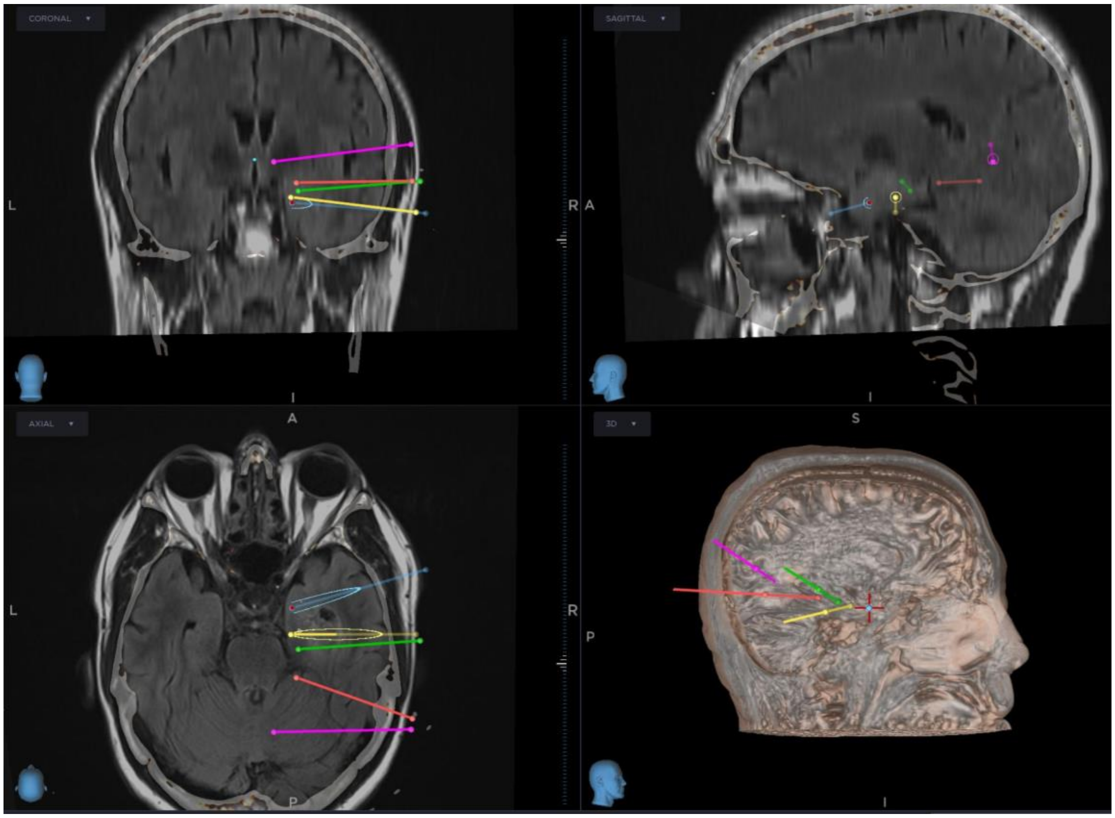


**Fig. S5. Intracerebral sEEG electrode locations in a patient with temporal lobe epilepsy**. Fusion of post-operative CT with pre-operative MRI showing contacts (colored lines) over tumor, amygdala, and hippocampus. For Surgical procedures, clinical management, and recordings - see Methods.

| | **Anteroposterior (AP)** | **Mediolateral (ML)** | **Dorsoventral (DV)** |
|---|---|---|---|
| Intracerebral | | | |
| *Medial Pre-Frontal cortex* | +3.2 | ±0.6 | −6 |
| *Hippocampus (CA1)* | -5.2 | -5.4** | -7.9 |
| *Lateral Entorhinal Cortex (LEC)* | -6.5 | ± 5.5 | -8.6 |
| *Visual cortex (V1)* | −5.25 | ± 3.40 | -1.8 |
| *Basolateral Amygdala (BLA)* | -2 | -4.6** | -9 |
| Epidural | | | |
| *Anterior Right (Epi. Ant. R.)* | -1 | -1.5** | X |
| *Posterior Right (Epi. Post. R.)* | -7 | -1.25** | X |
| Reference | | | |
| *Cerebellum* | *-10.35* | *-3.5** | X |

**Table 1: Implantation coordinates.**

All the coordinates in the AP axis are in reference to the Bregma and in mm.

* Left side only

** Right side only
